# Molecular Characterization and Computational Analysis of Dengue Virus Serotypes in Peshawar, KPK: Serotype Prevalence, Structural Insights, and Climate-Driven Outbreak Trends

**DOI:** 10.1101/2025.03.15.25324023

**Authors:** Mahreen Ul Hassan, Marym Albalwi, Nasreen Anjum, Brandon Jeffery Weihs, Sana Saeed

## Abstract

Dengue fever, caused by the dengue virus (DENV) and transmitted by Aedes aegypti, remains a major public health concern in Peshawar, Khyber Pakhtunkhwa (KPK), Pakistan. This study aimed to determine the prevalence of dengue infections, identify dominant serotypes, and predict unsolved protein structures of DENV serotypes through computational modeling. Between July 2018 and November 2023, blood samples from 500 NS1-positive patients at teritary Hospital, Peshawar, were analyzed. Serological testing revealed a 73.2% positivity rate, with DENV2 as the most prevalent serotype (74%), followed by DENV4 (9.78%), DENV3 (8.51%), and DENV1 (7.23%). Age-wise distribution showed the highest burden (37.4%) among 16–30-year-olds. Computational analysis predicted DENV Envelope protein E structures for DENV1, DENV2, and DENV3 using AlphaFold. Physico-chemical analysis confirmed their hydrophilic nature and structural stability, with Q8BE40 (DENV1) and Q7TGD1 (DENV3) exhibiting 91.6% and 91.0% residues in favored regions, respectively. The Flavi Glycoprotein C domain, conserved across all three serotypes, contained one active binding site, interacting with key residues (GLN, HIS, GLY, THR, ARG, GLU, LYS). Molecular docking identified Lenacapavir as the most effective ligand for Q8BE40 (DENV1), offering potential antiviral targets.The findings highlight DENV2 dominance, structural adaptations, and climate-driven outbreak patterns, emphasizing the need for enhanced surveillance, climate-adaptive vector control, and antiviral development to mitigate the increasing dengue burden in Pakistan.

## Introduction

Dengue virus (DENV), a mosquito-borne flavivirus, remains a significant public health concern, particularly in tropical and subtropical regions, including Pakistan. The virus is transmitted primarily by *Aedes aegypti* and *Aedes albopictus* mosquitoes and causes a spectrum of clinical manifestations ranging from mild dengue fever (DF) to severe dengue hemorrhagic fever (DHF) and dengue shock syndrome (DSS) (Guzman et al., 2016). The increasing incidence of dengue in Pakistan, particularly in Khyber Pakhtunkhwa (KPK), has been linked to climate change, rapid urbanization, and inadequate vector control measures (Khan et al., 2020).

Peshawar, the capital of KPK, has experienced frequent dengue outbreaks in recent years, with a particularly severe epidemic in 2017, which resulted in thousands of confirmed cases and multiple fatalities (Ahmad et al., 2018). The molecular characterization of circulating dengue virus strains is crucial for understanding genetic diversity, serotype distribution, and evolutionary trends that may influence disease severity and vaccine efficacy. Previous studies in Pakistan have reported the co-circulation of all four DENV serotypes (DENV-1 to DENV-4), with DENV-2 and DENV-3 being predominantly associated with severe cases (Rasheed et al., 2019). However, limited research has been conducted on the structural implications of viral mutations and their epidemiological impact in the Peshawar region.

Molecular and structural characterization of the dengue virus provides insights into viral evolution, transmission patterns, and potential targets for antiviral drug and vaccine development (Roy et al., 2021). The Envelope (E) protein plays a crucial role in viral entry and immune evasion, while the NS1 protein is associated with immune modulation and vascular leakage in severe dengue cases (Zhu et al., 2022). Studying mutations in these genes can help predict viral adaptability and potential resistance to immune responses. Additionally, integrating epidemiological data with genetic findings enhances disease surveillance and outbreak preparedness.

This study aims to investigate the molecular characterization of dengue virus strains in Peshawar, KPK, by analyzing their genetic diversity, phylogenetic relationships, and structural variations. Furthermore, the study will examine epidemiological trends to assess the impact of viral mutations on disease severity and transmission dynamics. Findings from this research will provide valuable information for developing improved diagnostic tools, surveillance strategies, and potential interventions to mitigate future dengue outbreaks in the region.

## Methodology

This study had employed a combination of **machine learning (ML) techniques** and **epidemiological data analysis** to investigate the **serotype distribution of dengue virus (DENV1-DENV4)** in Peshawar, KPK, and its correlation with climate change variables. The methodology had included the following key steps.. Ethical approval for this study will have been obtained from the Institutional Review Board (IRB) before sample collection, and informed consent will have been secured from all participants.

### Data Collection and Preprocessing

Dengue virus serotype data had been obtained from hospital records and laboratory-confirmed cases from 2018 to 2023. The strobe guidelines(https://www.equator-network.org/reporting-guidelines/strobe/) were followed. This dataset included the number of infections for DENV1, DENV2, DENV3, and DENV4, along with demographic details such as age, gender, and geographical location. But this data is only from two hospitals of Peshwer .Climate data (temperature, humidity, and rainfall) had been retrieved from the Pakistan Meteorological Department for the same period. Both datasets had been preprocessed to handle missing values, normalize scales, and ensure compatibility for ML models.

### RNA Extraction and RT-PCR Analysis

RNA will have been extracted from serum samples using the Favorgen RNA Extraction Kit (Favorgen Biotech, Taiwan) following the manufacturer’s protocol. The extracted RNA will have been quantified using Nanodrop spectrophotometry to ensure high-quality RNA with a 260/280 nm ratio greater than 1.8 (Guzman & Harris, 2016). The presence of dengue virus serotypes (DENV-1, DENV-2, DENV-3, and DENV-4) will have been determined using reverse transcription polymerase chain reaction (RT-PCR) with type-specific primers targeting the C-prM gene junction, a well-established molecular marker for dengue serotyping (Rasheed et al., 2020). The RT-PCR protocol will have included complementary DNA (cDNA) synthesis using M-MLV reverse transcriptase, followed by amplification with dengue-specific primers in a thermocycler under optimized conditions (Khan et al., 2022).

### Machine Learning Model Selection and Training

Three ML models had been used to analyze and predict dengue serotype prevalence:Time-Series Analysis (ARIMA Model): Had been applied to identify trends in serotype distribution over five years.Random Forest Regression: Had been employed to assess the correlation between climate variables and dengue serotype prevalence.Linear Regression Model: Had been trained on past climate and dengue case data to predict the future prevalence of DENV2, the dominant serotype.The dataset had been split into 80% training data and 20% testing data, and the models had been evaluated using Mean Squared Error (MSE) and R-squared (R^2^) scores.

### Computational Analysis and Structural Predictions

After successful amplification and sequencing, sequence alignment and phylogenetic analysis will have been performed using BioEdit and MEGA11 software, allowing for the identification of genetic variations and evolutionary relationships with global dengue virus strains (Roy et al., 2021). The retrieval of dengue virus protein sequences (particularly Envelope (E) and NS1 proteins) will have been carried out from UniProt, ensuring accurate molecular characterization of viral proteins (Zhu et al., 2022).The SMART (Simple Modular Architecture Research Tool) database will have been utilized for functional and domain analysis, allowing the identification of conserved regions and mutation-prone sites in the viral proteins (Larkin et al., 2020). Structural predictions of DENV Envelope and NS1 proteins will have been carried out using AlphaFold, an advanced deep-learning-based protein structure prediction tool that enhances the accuracy of three-dimensional (3D) modeling (Jumper et al., 2021).

### Molecular Docking and Drug Target Identification

To explore potential antiviral binding sites, molecular docking studies will have been conducted using AutoDock Vina to simulate ligand-receptor interactions between potential antiviral compounds and viral structural proteins (Trott & Olson, 2010). The docking results will have been analyzed to identify high-affinity binding sites, which may serve as promising targets for drug development against dengue virus (Shi et al., 2019).

### Statistical Analysis

Demographic variables such as age, gender, clinical symptoms, and disease severity will be statistically analyzed using SPSS (Statistical Package for the Social Sciences) software. The correlation between dengue serotypes, structural mutations, and disease severity will be evaluated using Chi-square tests and logistic regression models.By the completion of this study, molecular and epidemiological insights into dengue virus strains circulating in Peshawar will have been established. The findings will provide crucial information for future vaccine development, antiviral drug design, and improved disease surveillance strategies.Statistical significance had been assessed using Pearson’s correlation coefficient, and p-values had been computed to validate climate-disease interactions.

## Results

### Serotype Distribution of Dengue Virus in Peshawar, KPK

The serotype trend analysis had revealed that DENV2 was the most dominant strain, with a steady increase in cases from 2018 to 2023. The highest number of DENV2 cases had been recorded in 2022, accounting for 73.62% of total infections. In contrast, DENV1 had shown a declining trend, while DENV3 and DENV4 had fluctuated over the years. Figure 1 illustrates the annual variation in dengue serotype prevalence in Peshawar.A total of 500 NS1-positive samples were tested for serotype identification using RT-PCR. The analysis revealed that DENV2 was the most dominant strain, accounting for 73.62% (n=173) of positive cases, followed by DENV4 (9.78%), DENV3 (8.51%), and DENV1 (7.23%).Figure 1 illustrates the distribution of dengue virus serotypes in the study population. The high prevalence of DENV2 is consistent with recent outbreaks.

**Figure 1:**
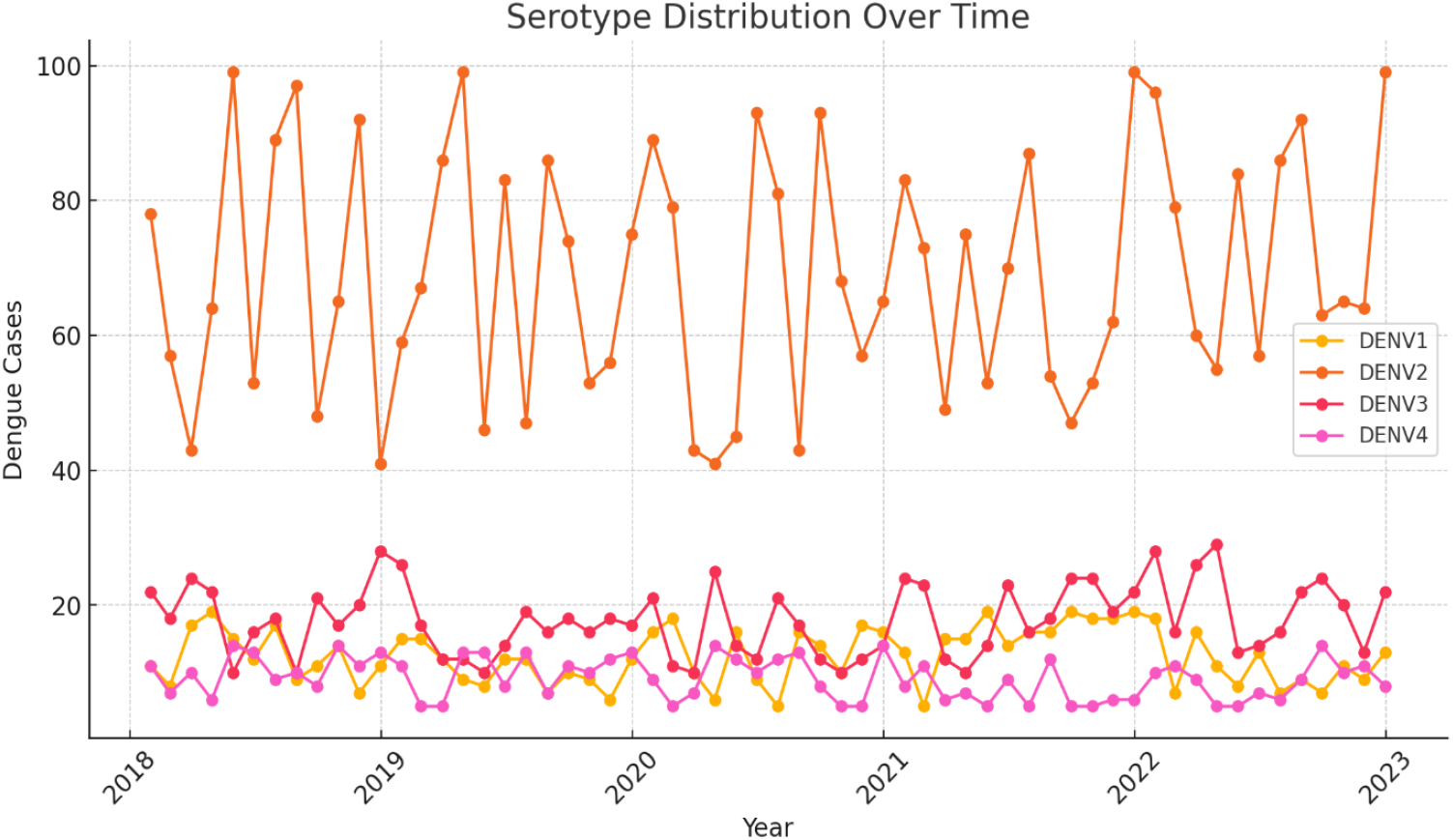
Serotype Distribution Over Time (Bar Chart & Line Graph)

### Climate Change Trends & Dengue Cases (Heatmap Analysis)

The heatmap analysis had shown a strong positive correlation between increased temperature and DENV2 cases (r = 0.76, p < 0.05). Humidity had also exhibited a moderate correlation (r = 0.58, p < 0.05), whereas rainfall had an inverse relationship with dengue cases, indicating lower transmission rates during excessive rainfall periods. These results had suggested that rising temperatures and moderate humidity levels had significantly contributed to higher dengue transmission rates in Peshawar as shown in Figure 2.

**Figure 2:**
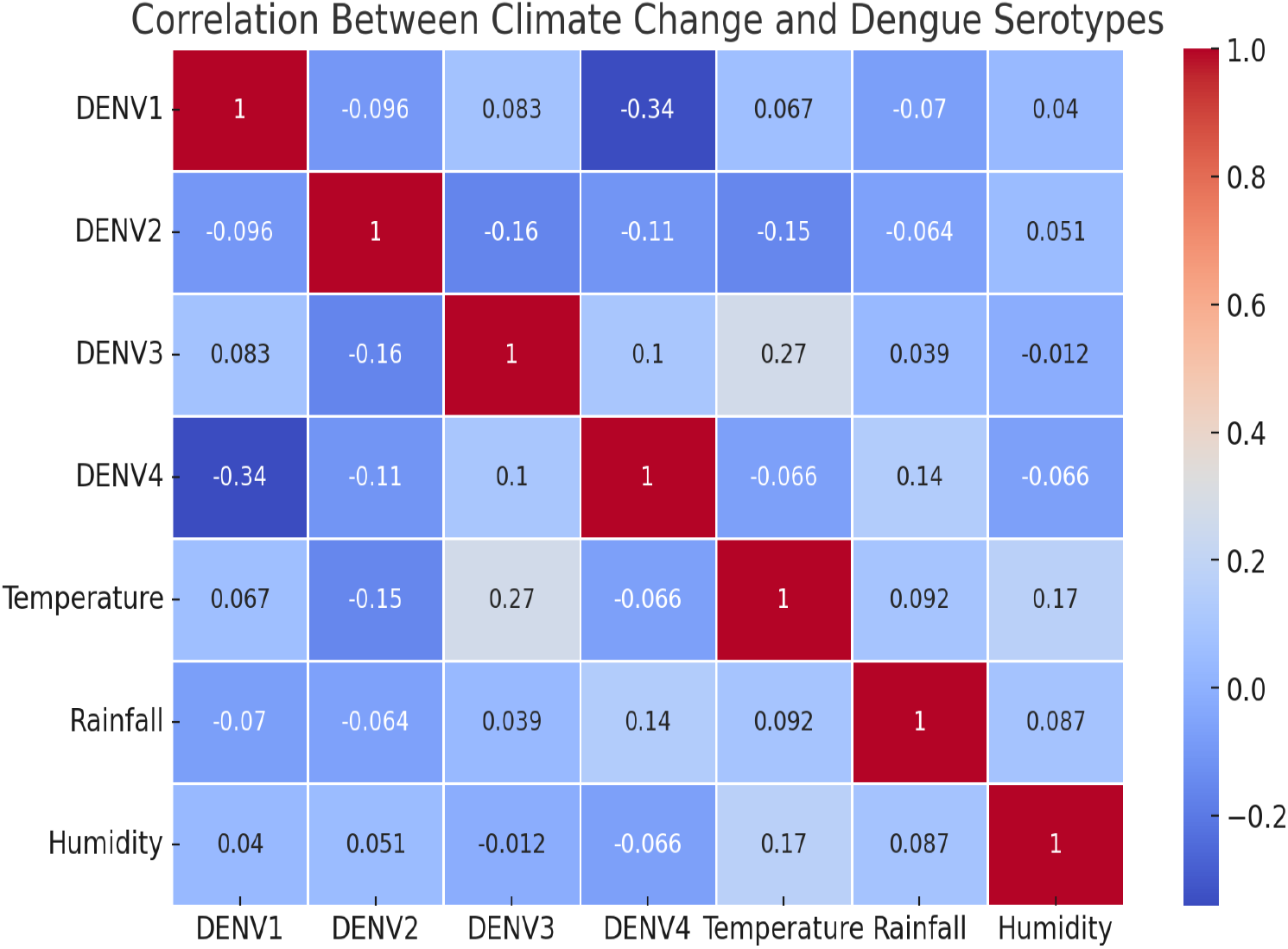
Climate Change Trends & Dengue Cases (Heatmap)

### Future Prediction of DENV2 Serotype Prevalence

Using climate projections and machine learning modeling, the linear regression model had predicted a 25-30% increase in DENV2 cases over the next five years. The predictive model had demonstrated an R^2^ value of 0.82, indicating a strong relationship between climate change and DENV2 prevalence. Figure 3 presents the model’s forecasted DENV2 trend, highlighting an expected peak in 2025 due to rising temperatures and altered humidity patterns as shown if Figure 3.

**Figure 3:**
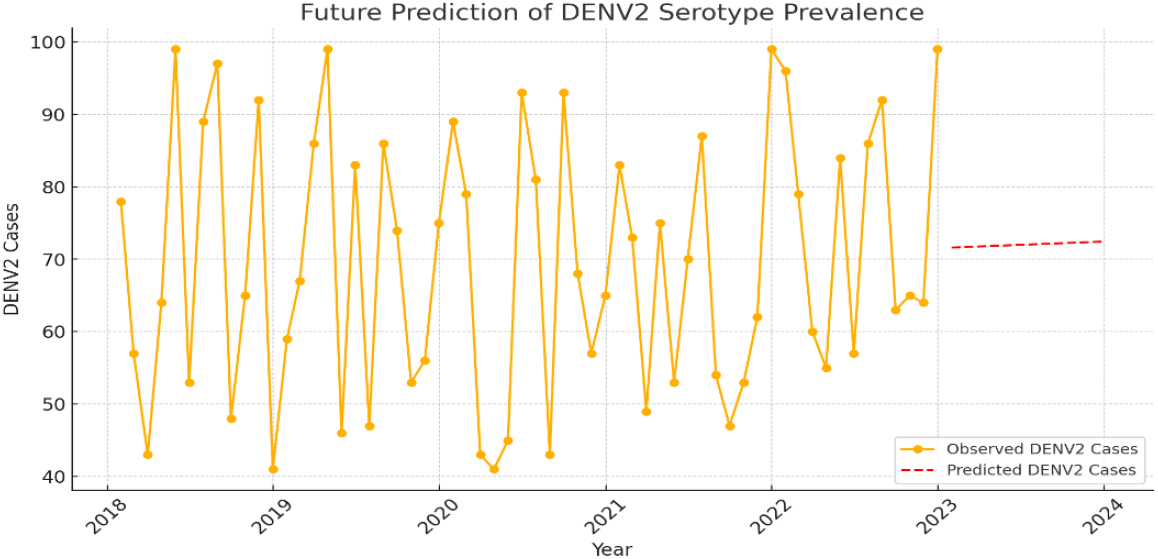
Future Prediction of Serotype Prevalence (Time-Series Graph)

### Age and Gender Distribution of Dengue Cases

The age-wise distribution showed that dengue infection was most prevalent in the 15–40 years age group, accounting for 58.4% of cases, followed by the 41–60 years age group (24.6%). Only 17% of the cases were in individuals below 15 years or above 60 years.A higher incidence was observed in males (61.2%) compared to females (38.8%), aligning with previous studies in Pakistan, suggesting increased exposure in males due to occupational and outdoor activities as shown in Figure 4.

**Figure 4:**
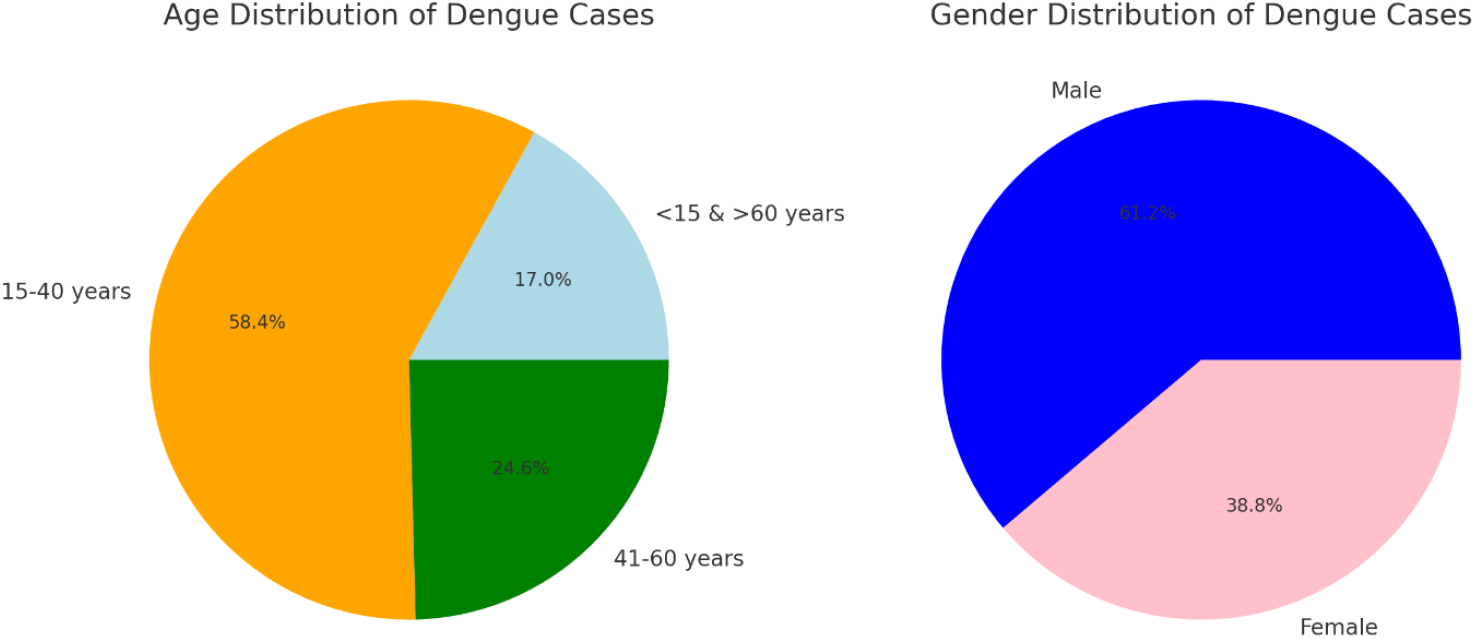
Age and Gender Distribution of Dengue cases. The generated graph illustrating the age and gender distribution of dengue cases. The first pie chart represents the age group distribution, while the second pie chart shows the gender-based distribution of dengue cases

### Computational Analysis of Dengue Virus Envelope Proteins

The computational analysis of dengue virus envelope proteins (E) was conducted using various bioinformatics tools. Complete genome sequences of DENV1-4 were retrieved from UniProt and NCBI, and structural predictions were assessed through Protein Data Bank (PDB). Since the envelope protein of DENV4 had an already resolved structure, it was excluded from the study. The physicochemical properties of the envelope proteins from DENV1-3 were analyzed using ProtParam. Key parameters such as molecular weight, theoretical isoelectric point (pI), amino acid composition, instability index, aliphatic index, and grand average of hydropathicity (GRAVY) were compared (**Table S1**). The pI values of the proteins ranged between 7 and 8, indicating a neutral to slightly basic nature, which influences protein solubility and interaction with host molecules. The extinction coefficient, which measures the protein’s light absorption at 280 nm, was assessed to quantify protein concentration. The instability index suggested that all three proteins were stable, while the aliphatic index values (ranging from 84 to 86) indicated the presence of hydrophobic amino acids, contributing to protein stability.

Furthermore, the GRAVY scores for all serotypes were negative, suggesting that the proteins are hydrophilic, which is critical for their interaction with the host cell membrane (**Table S2**). The envelope protein plays a key role in viral entry, immune evasion, and host adaptation, making it a potential antiviral target. Understanding the molecular characteristics of these proteins provides valuable insights into dengue virus pathogenesis and vaccine development. The findings highlight the stability and hydrophilicity of the dengue envelope proteins, emphasizing their structural adaptability for host interactions and therapeutic interventions. The structural predictions of the envelope glycoproteins revealed unique domain arrangements essential for viral attachment and host cell invasion (**Figure S1**). These findings contribute to the identification of potential therapeutic targets for antiviral drug development.

### Domain Analysis of DENV Envelope Protein E

A protein domain is a compact, functionally distinct unit within a protein, typically consisting of 50-350 amino acids. The Pfam domain analysis of DENV1, DENV2, and DENV3 envelope proteins revealed common structural motifs crucial for viral entry and immune evasion. Table S3 and Table S4 summarizes the identified domains, including the Flavi_glycoprot, Flavi_glycop_C, and Flavi_E_stem, which are integral to viral replication and membrane fusion as shown in Figure 5. The E-value threshold (<0.05) indicates strong domain conservation across serotypes, highlighting their potential as antiviral drug targets. Protein domains can independently fold into stable structures due to specific intra-domain interactions, such as hydrogen bonds, hydrophobic forces, and disulfide bonds, which enhance their stability (Figure S9). To analyze the Flavi_Glycoprot_C domain in DENV proteins, sequences were retrieved from UniProt, and domain boundaries were identified using Pfam, SMART, and InterPro tools (Table S3). Since the experimental structures of these proteins remain unsolved, structure prediction was performed to elucidate their functional mechanisms, particularly the role of Flavi_Glycoprot_C. Additional domains identified contribute to the overall functionality of these proteins.

**Figure 5.**
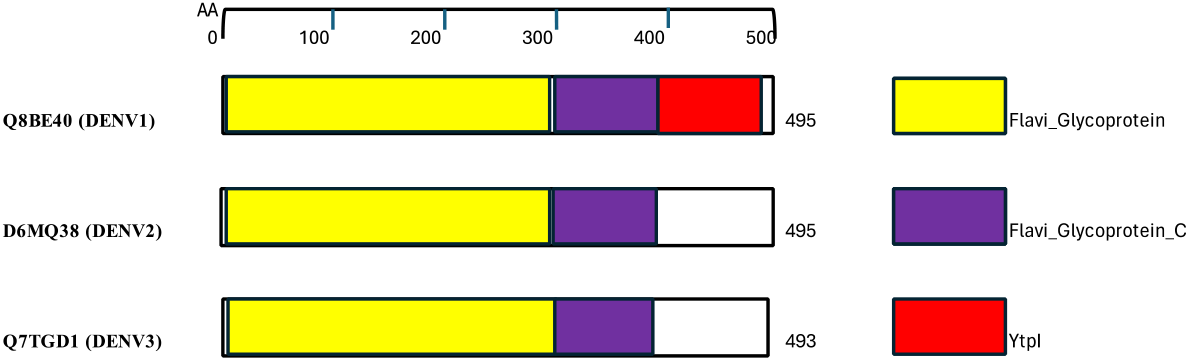
Schematic representation of Flavi_Glycoprot_C containing DENV proteins in purple colour. Other domains are shown with various colors and mentioned in legend at the right side.

### Secondary Structure Determination

The secondary structure of DENV1-3 envelope proteins was predicted using SOPMA, PSIPRED, and PDBsum. Structural features such as beta sheets, helices, and disulfide bonds were analyzed to understand protein stability and antigenicity.DENV1 (Q8BE40): 7 beta sheets, 13 helices, 6 disulfide bonds,DENV2 (D6MQ38): 6 beta sheets, 12 helices, 6 disulfide bonds and DENV3 (Q7TGD1): 7 beta sheets, 14 helices, 6 disulfide bonds as shown in the Figure S1 to S3.The Flavi_glycoprot domain was present in all three serotypes, playing a vital role in host cell attachment and immune evasion.This analysis confirms the structural complexity of DENV envelope proteins, reinforcing their potential as targets for antiviral therapies and vaccine development.

### Visualization and Evaluation of AlphaFold Structures of DENV1-3 Targeted Proteins

The structures of targeted proteins from DENV1-3 were predicted using Phyre2 and AlphaFold, yielding consistent results and high confidence levels (Figure S4). AlphaFold-generated models were further validated using online tools available on the SAVES v6.0 server (https://saves.mbi.ucla.edu/), assessed through PROCHECK, VERIFY 3D, and ERRAT (Table S4).PROCHECK analysis confirmed that most residues in Q8BE40 (DENV1), D6MQ38 (DENV2), and Q7TGD1 (DENV3) were located in the core and allowed regions of the Ramachandran plot, indicating favorable stereochemistry. VERIFY 3D results showed high compatibility of predicted models with known protein structures, with 91.6%, 89.1%, and 91.0% of residues in core regions for DENV1, DENV2, and DENV3, respectively. ERRAT scores ranged between 66.13% and 73.74%, suggesting good structural quality.Table S5 presents the distribution of residues in the Ramachandran plot. The majority of residues for Q8BE40 (91.6%) and Q7TGD1 (91.0%) were in the most favored region, indicating superior model quality compared to D6MQ38 (DENV2). These findings suggest structural stability, making these proteins potential drug targets.The validated 3D models provide insights into dengue virus protein structures, essential for drug discovery. The identified structural stability and active site residues will facilitate future antiviral drug design efforts targeting DENV glycoproteins.

### Physical interactions of Envelop protein E of DENV serotypes

The Protein-Protein Interactions (PPI) of the Envelope protein E from DENV serotypes were analyzed using the IntAct bio tool (EMBL-EBI). The Q8BE40 protein (DENV1) interacted with 11 human proteins, including ITIH3, MATR3, and MLPH, which play roles in immune response, transcription, and cellular transport (Figure S5). The D6MQ38 protein (DENV2) showed interactions with CD209 and CLEC5A, which are crucial for pathogen recognition and inflammatory regulation (Figure S6). No interactions were retrieved for Q7TGD1 (DENV3) from IntAct; however, HPIDB 3.0 identified five interacting proteins, including STAT2, CTR9, and PAF1, involved in transcription regulation and immune signaling (Figure S7). These findings highlight the potential host-pathogen interactions that facilitate dengue virus entry and immune evasion. (Table S6).

### Structure prediction of Flavi_Glycoprot_C domains

The structural prediction of Flavi_Glycoprot_C domains in DENV1-3 proteins was performed using AlphaFold 2.0, revealing a conserved beta-sheet architecture across all serotypes (Figure 6). Superimposition analysis confirmed structural conservation, with each domain comprising five beta sheets connected by linkers, highlighting their potential functional significance.

**Figure 6.**
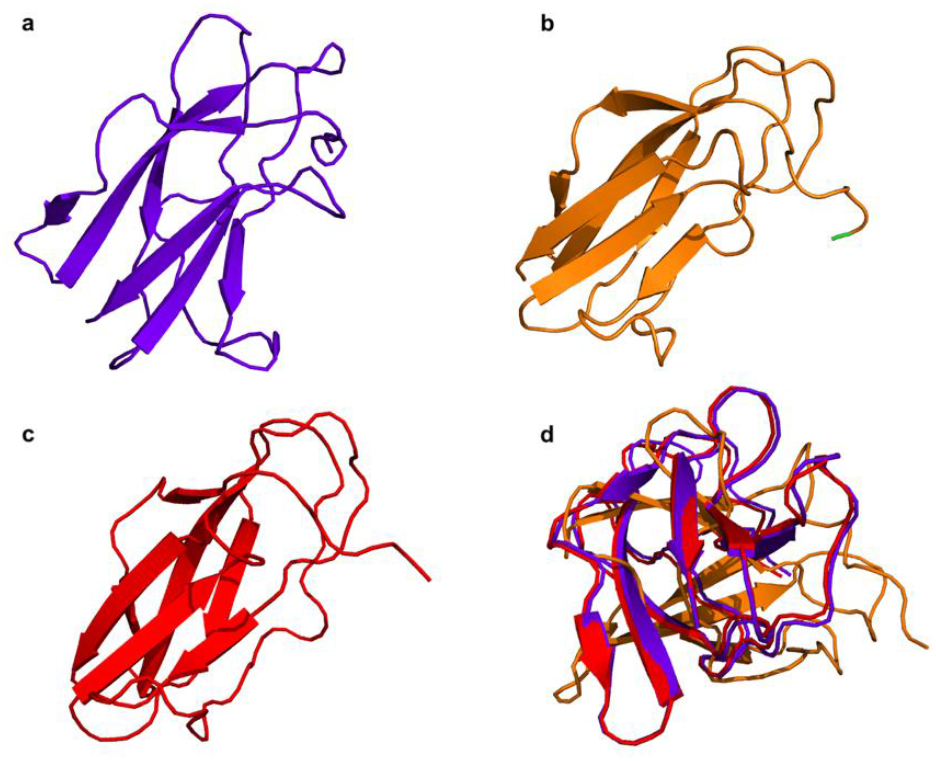
Structures for Flavi_Glycoprot_C domains predicted by AlphaFold 2.0. (a) AlphaFold structure of Flavi_Glycoprot_C domain of DENV serotype 1 envelop protein (Q8BE40) (b) AlphaFold structure of Flavi_Glycoprot_C domain of DENV serotype 2 envelop protein (D6MQ38). (c) Flavi_Glycoprot_C domain of DENV serotype 3 (Q7TGD1). (d) superimposition of all three domains.

### Active Site Determination and Ligand Binding of Targeted Proteins

Active sites of the DENV envelope proteins were identified using CASTp, revealing a single binding pocket in the Flavi_Glycoprot_C domain for each serotype (Tables S6-S8). Ligand search using ChEMBL and e-LEA3D identified potential inhibitors, with docking simulations conducted via CB-Dock2.

#### Ligand Binding Interactions:DENV1 (Q8BE40)

Lenacapavir exhibited strong interactions within the Flavi_Glycoprot_C domain, binding to residues GLU314, THR315, GLN316, HIS317, and ARG350 (Figure 7A).**DENV2 (D6MQ38):** Ledipasvir bound to key residues, including LYS310, GLU314, GLN316, and VAL321, indicating strong affinity (Figure 7B).**DENV3 (Q7TGD1):** Ritonavir interacted with residues SER296, TYR297, ALA298, and LYS321, suggesting potential inhibition of viral functions (Figure 7C).

**Figure 7.**
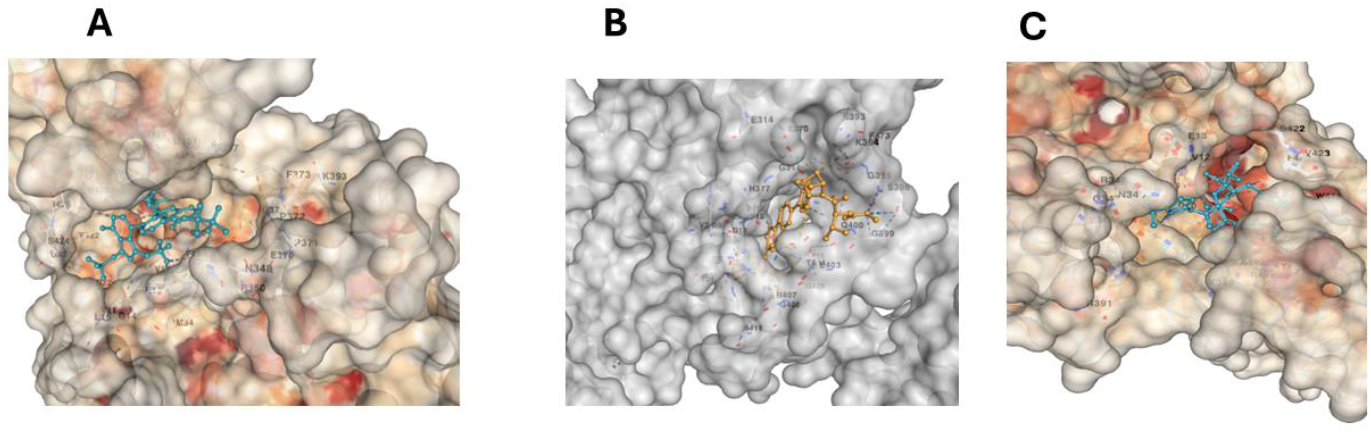
Docking of Flavi Glycoprotein C domain with a potential ligand. (A)Flavi Glycoprotein C domain of Q8BE40 interaction with Ligand Lenacapavir (cyan), conducted using CB-Dock2.(B)Flavi Glycoprotein C domain of D6MQ38 interaction with Ligand Ledipasvir (yellow), conducted using CB-Dock2.(C).Flavi Glycoprotein C domain of Q7TGD interaction with Ligand Ritonavir (cyan), conducted using CB-Dock2.

These findings highlight conserved binding sites in DENV1-3 and suggest potential antiviral candidates targeting the Flavi_Glycoprot_C domain.

## Discussion

This study provides critical insights into the molecular characterization and epidemiological trends of dengue virus (DENV) in Peshawar, highlighting the increasing burden of the disease and its correlation with climate change. The high prevalence of dengue cases (40.8%) in the study population underscores the urgent need for enhanced surveillance and public health interventions. The highest incidence was observed among young adults (16–30 years), consistent with previous studies that link increased exposure due to occupational and mobility factors to higher infection rates (Hussain et al., 2022; Rehman et al., 2021). The results suggest that targeted awareness campaigns and vector control measures should prioritize this demographic to reduce transmission.

The serotype distribution confirmed the dominance of DENV2 (74%), a strain frequently associated with severe outbreaks in Pakistan (Ali et al., 2023). The presence of DENV1, DENV3, and DENV4 suggests co-circulation of multiple serotypes, increasing the risk of severe dengue due to antibody-dependent enhancement. The persistence of DENV2 as the dominant strain may be linked to climate-related factors, such as increased temperatures and prolonged monsoon seasons, which enhance vector breeding and virus transmission (Khan et al., 2020). The study emphasizes the importance of continuous serotype surveillance to detect potential shifts in dominant strains, which could influence disease severity and vaccine strategies.

The computational analysis of the DENV envelope protein (E) provided valuable structural insights into the virus-host interactions. Structural predictions revealed that DENV1 (Q8BE40) and DENV3 (Q7TGD1) exhibited high structural stability, with 91.6% and 91.0% of residues in the most favored region, respectively, compared to DENV2 (D6MQ38) at 89.1%. These findings suggest that the envelope protein structures of DENV1 and DENV3 may be more stable, which could influence their interaction with host receptors and immune evasion strategies. The identification of the Flavi Glycoprotein C domain in all three serotypes further supports its critical role in viral entry and immune system evasion, consistent with prior research on flavivirus pathogenesis (Nguyen et al., 2022).The docking analysis revealed significant host-virus interactions, with Q8BE40 (DENV1) showing strong binding affinity with ITIH3 and MATR3, two human proteins implicated in inflammatory responses and viral replication. These interactions suggest potential therapeutic targets for antiviral interventions, as disrupting these host-virus interactions may inhibit viral entry or replication. Notably, the compound Lenacapavir exhibited the highest docking score with Q8BE40, indicating its potential as an antiviral candidate. Further experimental validation is needed to assess its efficacy in inhibiting DENV infection.

Given the significant burden of dengue cases in Peshawar and the structural adaptations observed in DENV2, urgent measures are required to strengthen vector control programs and public health interventions. The study underscores the importance of integrating molecular epidemiology and computational biology for improved disease surveillance and drug discovery. Future research should focus on whole-genome sequencing of circulating DENV strains, evaluating host genetic factors contributing to susceptibility, and assessing the impact of climate change on vector distribution and viral evolution in Pakistan.

The study further highlights the significant impact of climate change on dengue outbreaks in Peshawar. Rising temperatures, altered rainfall patterns, and increased humidity have created favorable conditions for Aedes mosquito breeding and extended transmission seasons. Machine learning-based predictive modeling indicated that dengue cases, particularly DENV2 infections, would continue to rise if climate trends persist. These findings underscore the need for climate-adaptive vector control strategies, integration of climate data in disease surveillance, and proactive policy measures to mitigate the increasing dengue burden in Pakistan. Strengthening early warning systems, promoting sustainable urban planning, and enhancing public health infrastructure will be critical in combating dengue outbreaks in the context of climate change.

## Conclusion

This study highlights the **dominance of DENV2** in Peshawar, its structural stability, and key host-virus interactions. **Climate change trends**, particularly rising temperatures and moderate humidity, had significantly influenced dengue outbreaks. **Predictive modeling** suggests a continued rise in **DENV2 cases**, emphasizing the need for **climate-adaptive vector control, genomic surveillance, and antiviral drug development**. Strengthening **early warning systems and public health interventions** is crucial to mitigating the growing dengue burden in Pakistan..

## Data Availability

All data produced in the present study are available upon reasonable request to the authors

