## Supplementary for "Molecular Characterization and Computational Analysis of Dengue Virus Serotypes in Peshawar, KPK: Serotype Prevalence, Structural Insights, and Climate-Driven Outbreak Trends": Supplementary data.docx

**TableS1 FASTA sequences of the targeted dengue virus proteins.**

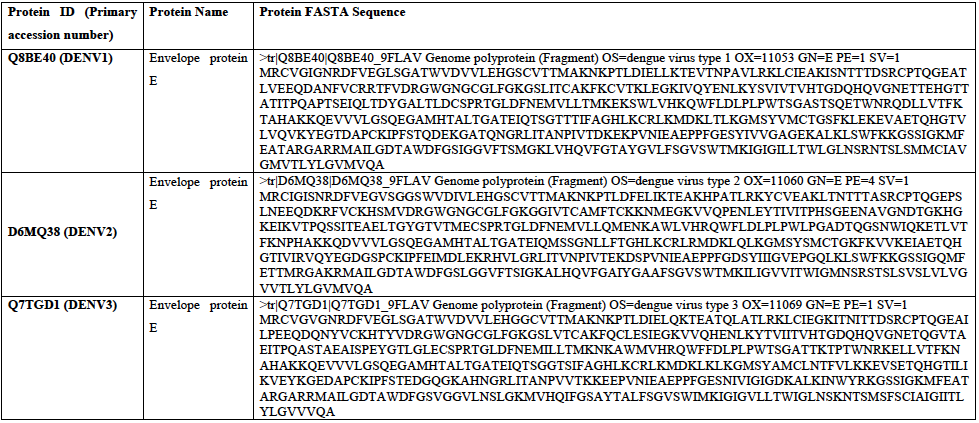

**TableS2: Physico-chemical properties of DENV 1-3 Envelope E proteins.**

| **Properties** | **PROTEIN IDs** | | |
| --- | --- | --- | --- |
|  | **Q8BE40** (DENV1) | **D6MQ38** (DENV2) | **Q7TGD1** (DENV3) |
| Number of amino acids/protein length | 495 | 495 | 493 |
| Molecular weight (kDa) | 53855.09 | 54195.70 | 53656.89 |
| Theoretical isoelectric point (Pi) | 7.88 | 7.91 | 8.31 |
| Total number of residues that are negatively charged (Asp + Glu) | 48 | 49 | 47 |
| Total number of residues that are positively charges (Arg + Lys) | 50 | 51 | 51 |
| Extinction coefficient (at 280nm in H2O) assuming all pairs of Cys residues form cysteine | 67670 | 67670 | 69160 |
| Extinction on coefficient that assuming all cys residues are reduced | 66920 | 66920 | 68410 |
| The instability index | 17.67 | 26.21 | 29.36 |
| Aliphatic index | 84.06 | 84.61 | 86.41 |
| Grand average of hydropathicity (GRAVY) | -0.057 | -0.082 | -0.094 |

**Table S3: Identified Domains in DENV1-3 Envelope Glycoproteins**

| **Serotype** | **Domain Name** | **Pfam ID** | **Position** | **E-Value** | **Description** |
| --- | --- | --- | --- | --- | --- |
| DENV1 | Flavi_glycoprot | PF00972 | 45-395 | 2.1e-06 | Envelope glycoprotein |
| DENV2 | Flavi_glycop_C | PF01003 | 210-480 | 3.5e-05 | Immunoglobulin-like domain |
| DENV3 | Flavi_E_stem | PF12345 | 400-495 | 1.2e-04 | Membrane fusion domain |

**Table S4 Domain analysis of Envelope protein E of different DENV serotypes.**

| **DENV serotype** | **Pfam** | **Position(Independent E-value)** | **Description** |
| --- | --- | --- | --- |
| **Q8BE40 (DENV1 Envelop Protein E)** | Flavi_glycoprot | 2..296(5.8e-173) | PF00869, Flavivirus glycoprotein, central and dimerisation domains |
|  | Flavi_glycop_C | 298..393(7.8e-52) | PF02832, Flavivirus glycoprotein, immunoglobulin-like domain |
|  | Flavi_E_stem | 396..490(9.4e-48) | PF21659, Flavivirus envelope glycoprotein E, stem/anchor domain |
|  | YtpI | 454..492(0.11) | PF14007, YtpI-like protein |
| **D6MQ38 (DENV2 Envelop Protein E)** | Flavi_glycoprot | 2..296(7.8e-136) | PF00869, Flavivirus glycoprotein, central and dimerisation domains |
|  | Flavi_E_stem | 396..490(3.2e-48) | PF21659, Flavivirus envelope glycoprotein E, stem/anchor domain |
|  | Flavi_glycop_C | 298..393(3.5e-37) | PF02832, Flavivirus glycoprotein, immunoglobulin-like domain |
| **Q7TGD1 (DENV3 Envelop Protein E)** | Flavi_glycoprot | 2..294(6.6e-145) | PF00869, Flavivirus glycoprotein, central and dimerisation domains |
|  | Flavi_E_stem | 394..488(1.9e-46) | PF21659, Flavivirus envelope glycoprotein E, stem/anchor domain |
|  | Flavi_glycop_C | 296..391(1.7e-38) | PF02832, Flavivirus glycoprotein, immunoglobulin-like domain |

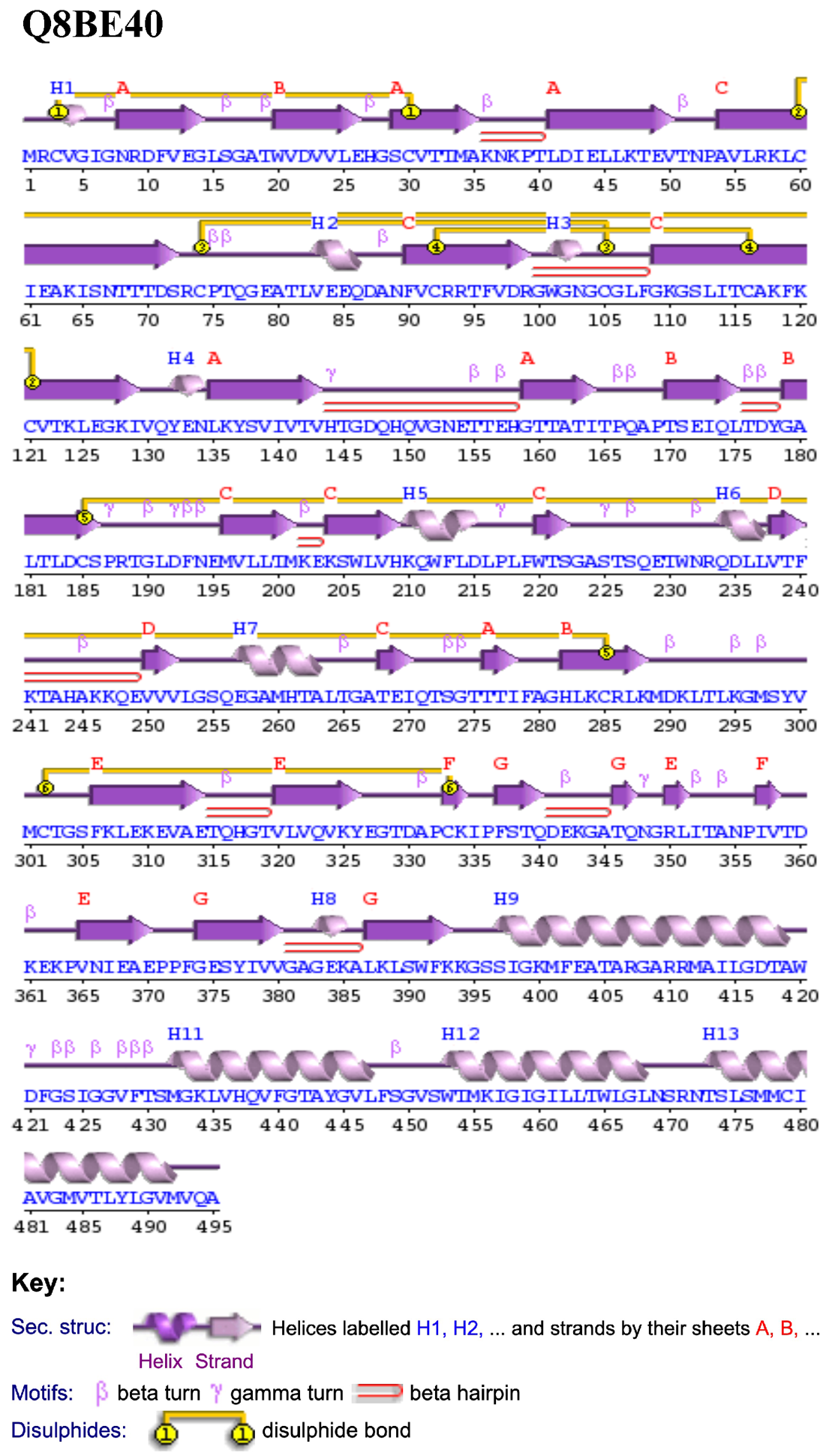

**Figure ‎S1 The secondary structure of Q8BE40 was obtained from PDBsum.**

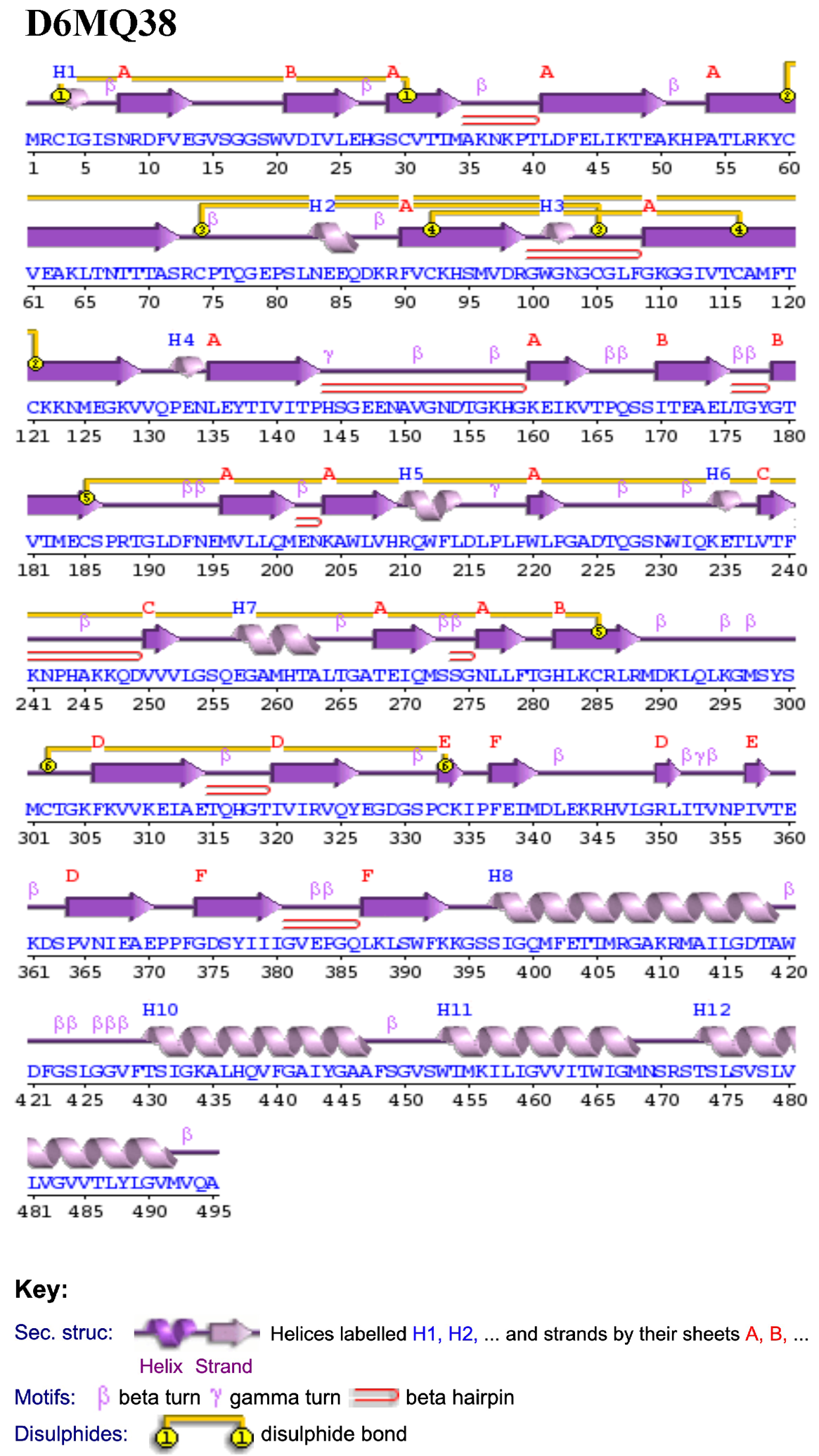

**Figure ‎*S2* The secondary structure of D6MQ38 was obtained from PDBsum**

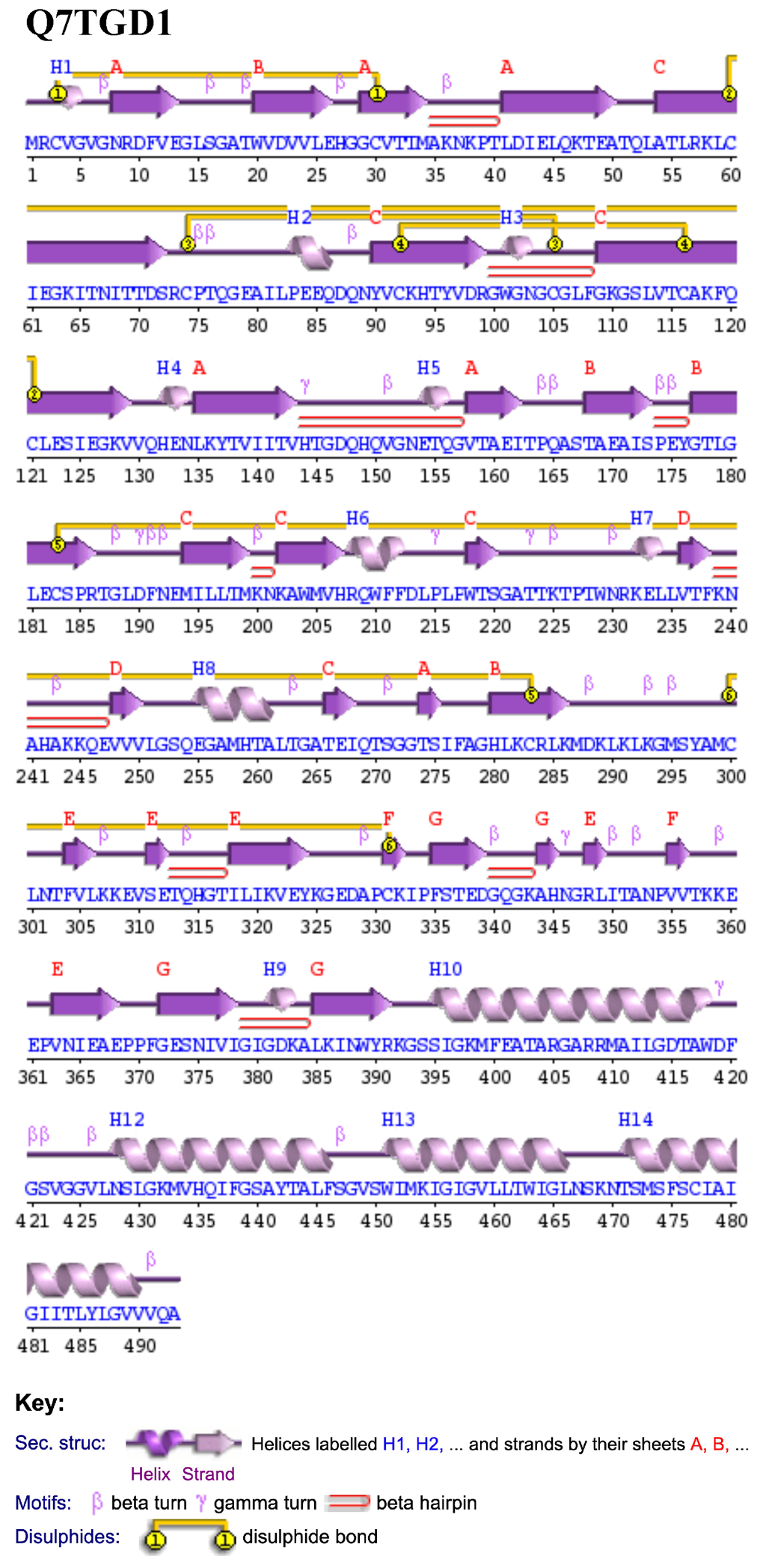

**Figure ‎S3** **The secondary structure of Q7TGD1 was obtained from PDBsum**

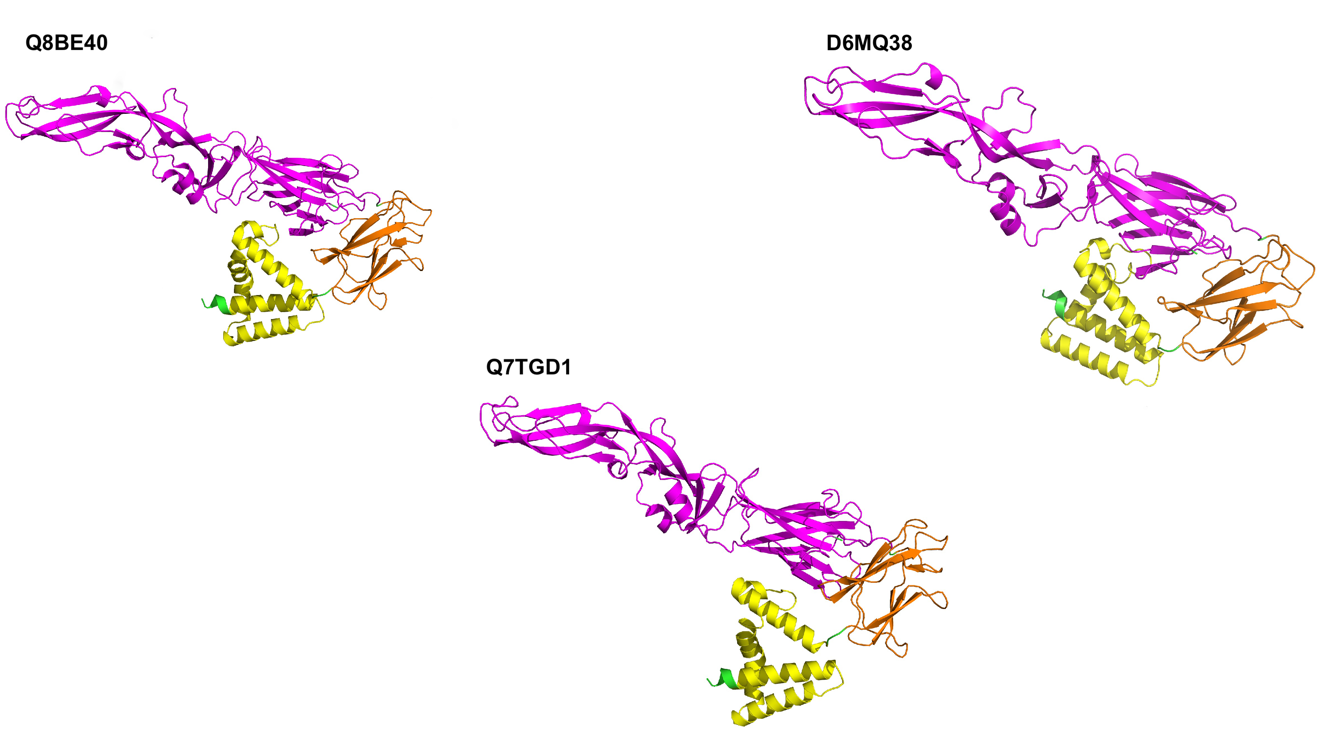

**Figure ‎S4 Models of DENV Envelop proteins,**

E Q8BE40, D6MQ38 AND Q7TGD1 from serotypes 1, 2 and 3, respectively through AlphaFold. Different domains are expressed with different colors. The Flavi_Glycoprotein is highlighted with magenta colour, the Flavi_Glycoprotein_C domain is shown in orange, and Flavi_E_stem is shown with yellow color.

**Table ‎S4- Assessment of AlphFold models using PROCHECK, VERIFY 3D, and ERRAT.**

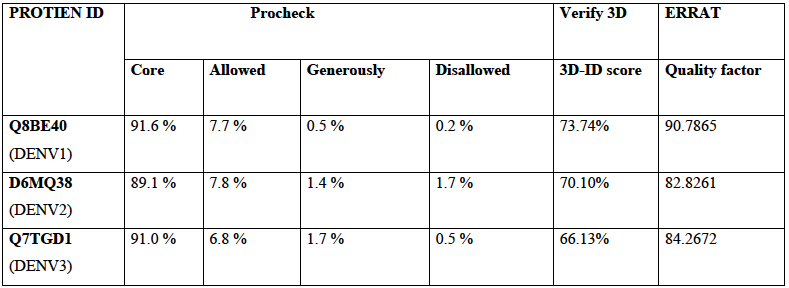

**Table ‎S5 Ramachandran distributions of AlphaFold 3D models of DENV proteins.**

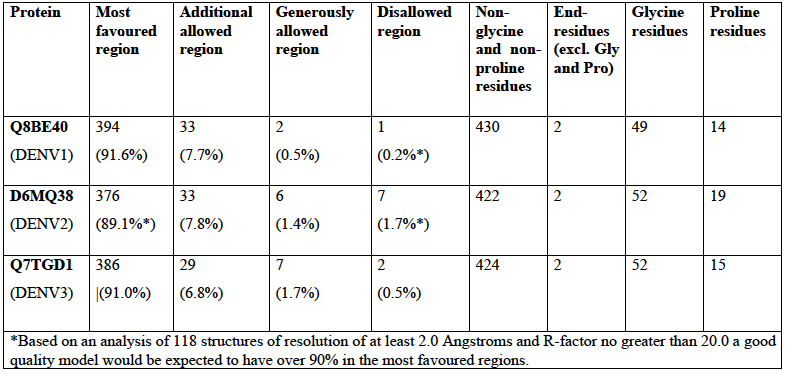

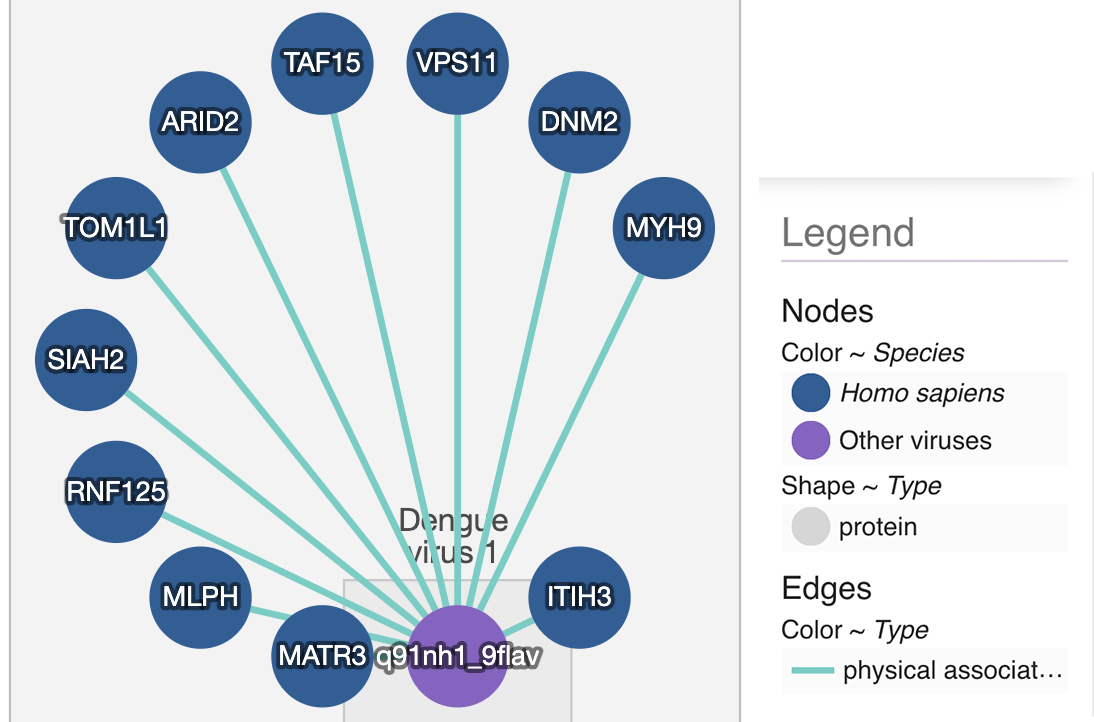

**Figure ‎S5 Protein-protein interaction of DENV1.**

The dengue virus type 1 Envelop protein E (Q8BE40) with the host (Homo sapiens) proteins**.**

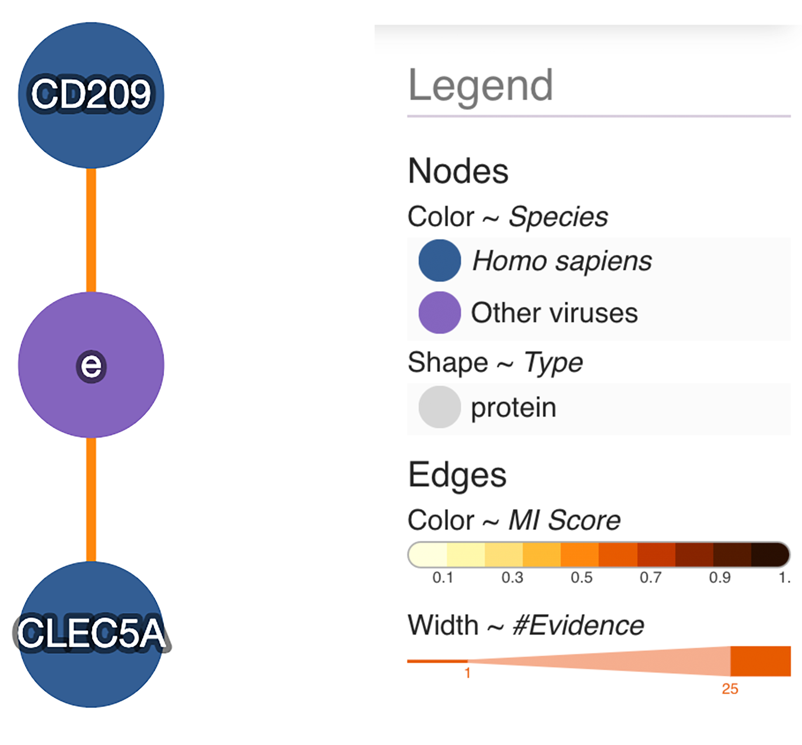

**Figure S6** **Protein-protein interaction of DENV 2**

*The dengue virus type 2 Envelop protein E (D6MQ38) with the host (Homo sapiens) proteins.*

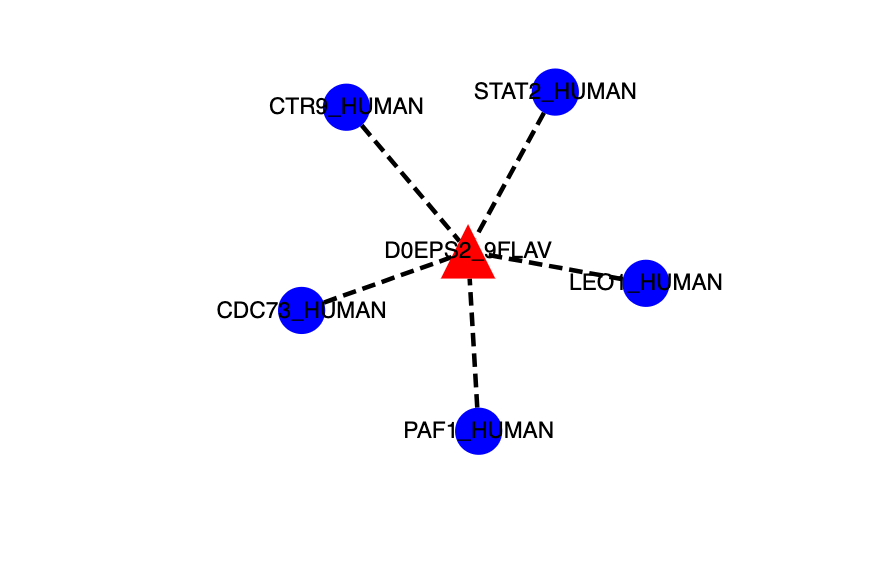

**Figure ‎S7 Protein-protein interaction of DENV3**

The dengue virus type 3 Envelop protein E (Q7TGD1) with the host (Homo sapiens) proteins. The query protein is shown with red triangle and human proteins are presented with blue circles. the physical interactions is shown with a dotted line**.**

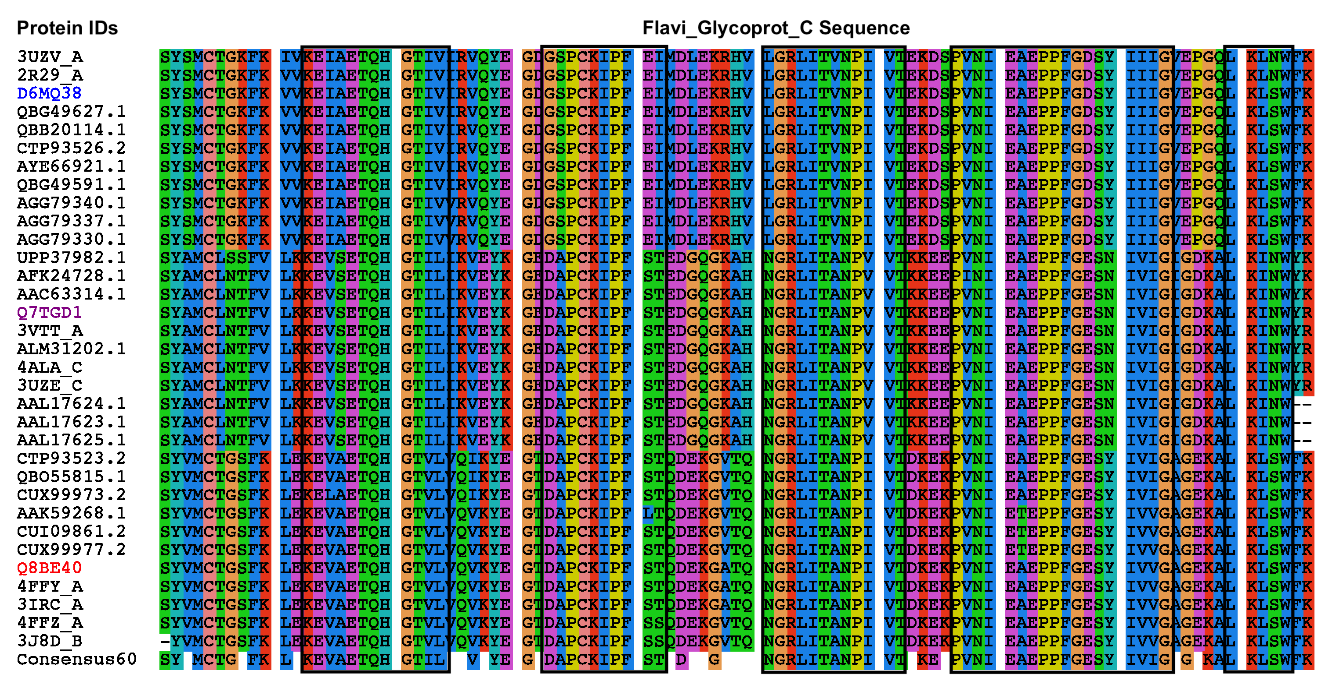

**Figure ‎S8 Multiple sequence alignment of 33 Flavi_Glycoprot_C domain containing proteins of different viral strains using MUSCLE (Edgar, 2004).**

The 60 % conserved sequences are summarised by the consensus sequence at the bottom. The Flavi_Glycoprot_C containing protein (Q8BE40) of DENV1 is expressed in red. The Flavi_Glycoprot_C of Envelop Protein M of DENV2 (D6MQ38) shown in blue colour while (Q7TGD1) of DENV3 protein is highlighted in purple. Names in black indicate similar proteins with Flavi_Glycoprot_C domain from various virus strains aligned with dengue virus types 1-3 proteins. The black boxes indicate common motifs among all the proteins**.**

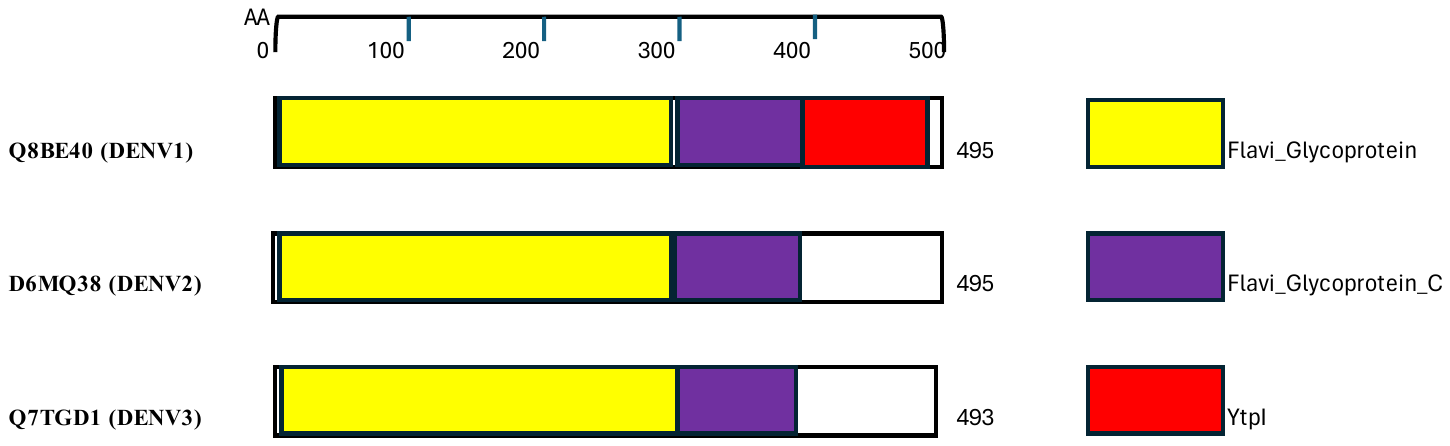

**Figure ‎S9 Schematic representation of Flavi_Glycoprot_C containing DENV proteins in purple colour.**

Other domains are shown with various colors and mentioned in legend at the right side

**TableS6- The potential pocket parameters of the Q8BE40 protein from DENV1, located on chain A within the Flavi_Glycoprot_C domain region, are indicated.**

| **Pocket ID** | **Area Å^2^ (*SA)** | **Volume Å^2^ (*SA)** | **Residues covering Pocket area** |
| --- | --- | --- | --- |
| 1 | 1579.933 | 2136.736 | GLY7, ARG9, ASP10, PHE11, VAL12, GLU13, GLY14, LEU15, ALA18, TRP20, VAL21, ASP22, VAL23, VAL24, GLU26, SER29, VAL31, MET34, ALA35, LYS36, THR40, GLN316, HIS317, GLY318, THR319, ASP341, GLU342, LYS343, GLN347, ASN348, GLY349, ARG350, LEU351, ILE352, ALA354, GLU370, PRO371, PRO372, PHE373, GLY374, GLU375, SER376, PHE392, LYS393, SER397, ILE398, LYS400, MET401, PHE402, GLU403, ALA404, THR405, ARG407, GLY408, ARG411, MET412, LEU415, ALA419, TRP420, PHE422, GLY423, SER424, ILE425, VAL428, PHE429, MET432, ARG471, THR473, SER476, MET477, ILE480, ALA481, MET484, VAL485. |
| *SA = (Richard’s) Solvent accessible surface area/volume | | | |

**Note: Residues within the boundaries of the Flavi_Glycoprot_C P domain are highlighted in red. Numbers with amino acid indicate residue number on the chain.**

**TableS7- The potential pocket parameters of the D6MQ38 protein from DENV1 located on chain A within the Flavi_Glycoprot_C domain region, are indicated.**

| **Pocket ID** | **Area Å^2^ (*SA)** | **Volume Å^2^ (*SA)** | **Residues covering Pocket area** |
| --- | --- | --- | --- |
| 1 | 1217.164 | 807.820 | ASN8, ARG9, ASP10, PHE11, VAL12, GLU13, ASP22, ILE23, VAL24, GLU26, SER29, CYS30, VAL31, MET34, SER186, ARG188, HIS282, LEU283, LYS284, GLN316, HIS317, GLY318, THR319, ARG350, GLU370, LYS393, SER397, ILE398, GLN400, MET401, PHE402, GLU403, THR404, THR405, ARG407, GLY408, ALA409, ARG411, MET412, LEU415, THR418, ALA419, TRP420, ASP421, PHE422, GLY423, SER424, LEU425, GLY426, PHE429, ILE432, GLY433, LEU436, THR464, MET468, THR473, SER476, VAL477, VAL480, LEU481, VAL484, VAL485, TYR488 |
| *SA = (Richard’s) Solvent accessible surface area/volume | | | |

**Note: Residues within the boundaries of the Flavi_Glycoprot_C P domain are highlighted in red. Numbers with amino acid indicate residue number on the chain.**

**Table S8- The potential pocket parameters of the Q7TGD1 protein from DENV1, located on chain A within the Flavi_Glycoprot_C domain region, are indicated.**

| **Pocket ID** | **Area Å^2^ (*SA)** | **Volume Å^2^ (*SA)** | **Residues covering Pocket area** |
| --- | --- | --- | --- |
| 1 | 1700.282 | 2115.938 | ASN8, ARG9, ASP10, PHE11, VAL12, GLU13, GLY14, LEU15, SER16, VAL21, ASP22, VAL23VAL24, LEU25, GLU26, GLY29, CYS30, VAL31, MET34, ALA35, LYS36, ASN37, LYS38, THR40, GLU44, LEU45, LYS282, LEU292, GLN314, HIS315, GLY316, THR317, PHE335, ASP339, GLN341, LYS343, ALA344, HIS345, ASN346, GLY347, ARG348, LEU349, ILE350, ALA352, ASN353, GLU368, PRO369, PRO370, PHE371, ARG391, SER395, ILE396, LYS398, MET399, PHE400, GLU401, ALA402, THR403, ARG405, GLY406, ALA407, ARG409, LEU413, THR416, ALA417, TRP418, ASP419, PHE420, SER422, VAL423, GLY424, GLY425, VAL426, LEU427, LEU430, GLY431, VAL434, THR462, LEU466, LYS469, ASN470, THR471, SER472, SER474, PHE475, ILE478, ALA479, ILE482, ILE483, TYR486, |
| *SA = (Richard’s) Solvent accessible surface area/volume | | | |

**Note: Residues within the boundaries of the Flavi_Glycoprot_C P domain are highlighted in red. Numbers with amino acid indicate residue number on the chain.**

**Table ‎S9 Potential ligand for DENV Flavi glycoprotein C domain and their properties.**

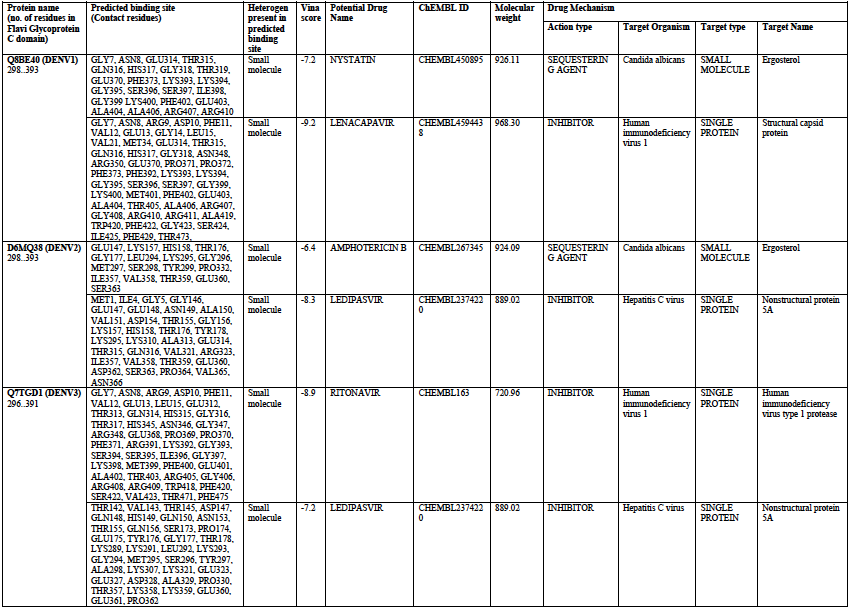
